# The Effect of COVID-19 Public Health Measures On Mental Health in California

**DOI:** 10.1101/2023.04.17.23288661

**Authors:** Milad Asgari Mehrabadi, Erika L. Nurmi, Jessica L. Borelli, Natalie Lambert, Amir M. Rahmani, Charles A. Downs, Melissa D. Pinto

## Abstract

California was the first state to implement statewide public health measures, including lockdown and curfews, to mitigate transmission of SARS-CoV-2. The implementation of these public health measures may have had unintended consequences related to mental health for persons in California. This study is a retrospective review of electronic health records of patients who sought care in the University of California Health System to examine changes in mental health status during the pandemic. Data were extracted prior to the pandemic (March-October 2019) and during the pandemic (March-October 2020). Weekly values of new mental health disorders were extracted and further classified based on age. Paired t-tests were performed to test for differences in the occurrence of each mental health disorder for each age group. A two-way ANOVA was performed to assess for between group differences. When compared with pre-pandemic diagnoses, persons aged 26-35 had the greatest increase in mental health diagnoses overall during the pandemic, specifically for anxiety, bipolar disorder, depression, mood disturbance, and psychosis. The mental health of persons age 25-35 were more affected than any other age group.

## Introduction

There have been more than 9 million cases, equating to over 90,000 deaths, of severe acute respiratory syndrome coronavirus 2 (SARS-CoV-2) infection in California since December of 2019^1^. In an effort to prevent viral transmission, California was the first state to implement a statewide public health measures, which included lockdown and curfews. Social isolation, economic concerns, grief, fear and the loss of loved ones in addition to the uncertainty of the pandemic and SARS-CoV-2 contributed to an increase in stress amongst the general public. Although the reason for the implementation of public health measures was to protect the public from viral transmission, it is likely such measures had unintended consequences for mental health, specifically, new diagnoses of mental health disorders: anxiety, bipolar disorder, post-traumatic stress disorder, depression, selfharm, suicide, and psychosis, among others. Because California uniformly enacted public health measures at the side-wide level, data throughout the state can be used to describe the potential influence of the environment on mental health.

There is a compelling body of literature to support the role of the environment: either protective or harmful to one’s mental health ^2^. When human developmental stage is considered, there are critical periods–especially during childhood and adolescence– in which the environment is highly influential in shaping a young person’s mental health, current and well into the future. Similarly, it is well-documented among the elderly, that social isolation is detrimental to health. Thus, it is likely that public health measured did affect all age groups.

The purpose of this study was to examine the influence of the COVID-19 pandemic, including the implementation of public health measures, on mental health disorders across the lifespan and between different age groups. This retrospective review uses data, collected before and after the start of the pandemic, from the University of California COVID Research DataSet (UC-CORDS) to determine the frequency of mental health issues across the lifespan during these time frame

## Method

### Sample

Data were extracted from the medical records within the University of California Healthcare System at two time points: prior to (March-October of 2019) and during the pandemic (March-October 2020). Data were examined to between the two time periods to determine any difference in new diagnoses of mental health problems across the lifespan. The data set contains comprehensive, structured information on patients admitted to the hospitals at the University of California Health’s five academic health centers (i.e., UC

Davis Health, UC San Diego Health, UC Irvine Health, UCLA Health, and UCSF Health). This data set provides a wide range of information, including COVID-19 test results, and weekly values of new mental health disorders diagnosed by UC-Health hospitals. It does not necessarily contain unique subjects since one subject that developed anxiety and bipolar is recorded twice – one for anxiety and one for bipolar. Weekly values of new diagnoses of mental health disorders were extracted from UC-CORDS and further classified based on age (less than 19 (*<* 19), between 19 and 25 (19-25), 26-35, 36-45, 46-55, 56-65, 66-75, 76-85 and above 85 (*>* 85) years old).

The UC-CORDS data set contains different diagnoses coded using SNOMED vocabulary ^3^. For analysis purposes, we have categorized various mental health disorders into 12 bins. *Bipolar, post-traumatic stress disorder (PTSD), depression, mood, psychosis, self-harm, suicide, personality*, and *anxiety* are straightforward. *Child/adolescent behavioral* contains disorders related to adolescents, such as hyperactivity disorder. *Substance-induced mental health* consists of the disorders developed due to the use of substances of abuse or medications (e.g., delusional disorders); however, the disorders in usage are categorized as *Substance use*.

### Analysis

Statistical analyses were performed using R (version 4.0.3). Paired t-tests were performed to test differences in the occurrence of each mental health disorder for each age group. A two way ANOVA was performed to assess for between group differences. A *P* value of *<* 0.05 was considered significant.

## Results

The occurrence of new diagnoses of mental health disorders for 2019 (pre-pandemic) and 2020 (during the pandemic) based on age are reported in Table 1. There was a significant change in mental health diagnoses observed for specific groups (*<* 19, 19-25, 26-35, 56-65, 76-85). The largest increase in mental health diagnoses (overall) was observed in the 26-35 age group (*P <* .001), See Fig 1a.

**Table 1.**
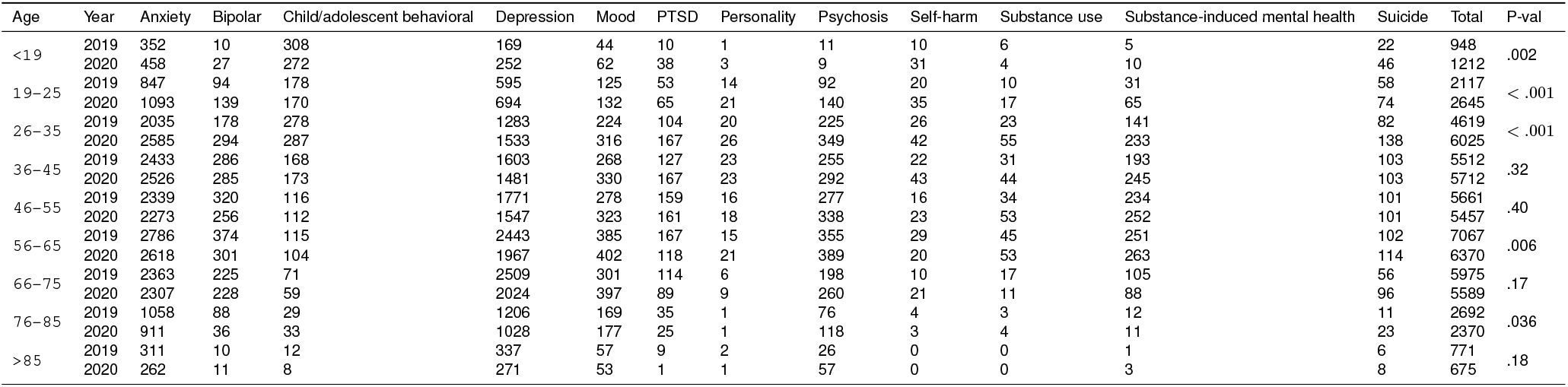
Reported values of each disorder per age group in 2019 and 2020.

**Figure 1.**
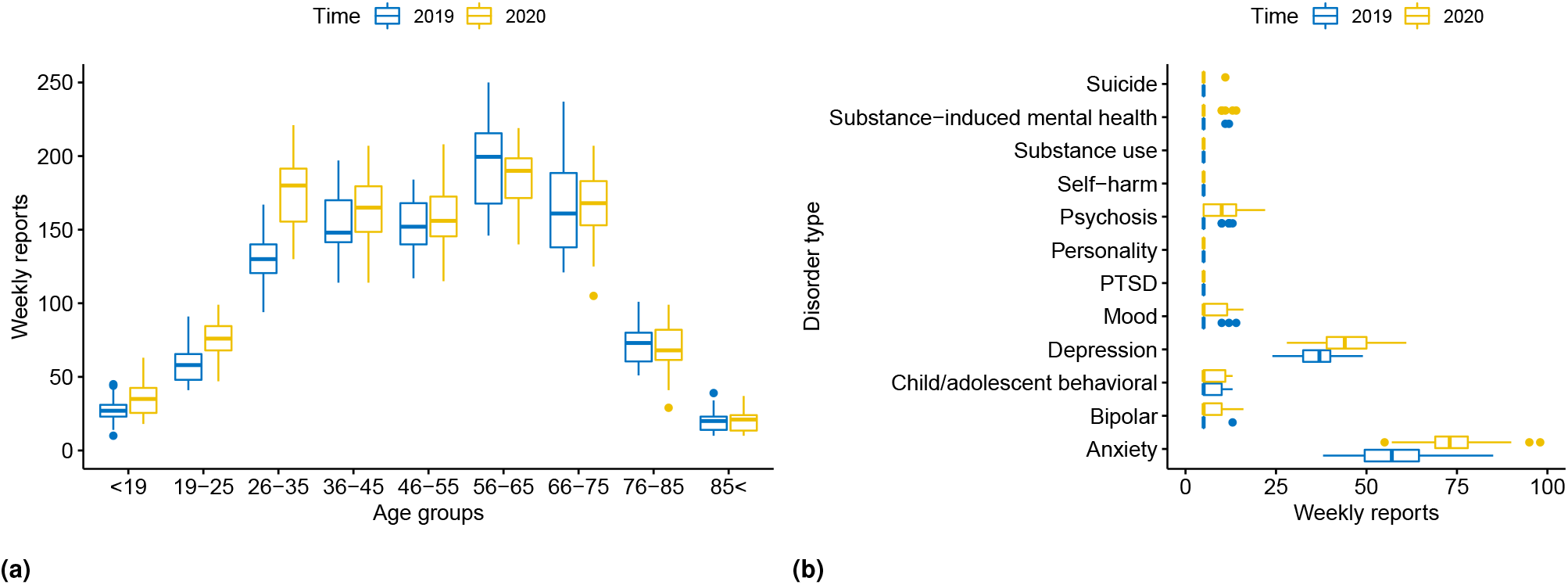
Distribution of total mental health disorder reports per age group (1a) and per each disorder for 26-35 years old population (1b).

In addition, the two-way ANOVA test for 2019 and 2020 showed significant differences in age groups over time (*P <* .001 and *P* = .004 respectively), justifying different age groups with different intercepts and slopes in both 2019 and 2020.

Since the most significant changes were observed for age groups 26-35, we examined specific mental health disorders by type in this age group using paired t-tests. There was a significant increase in anxiety (*P <* .001), bipolar (*P* = .02), depression (*P <* .001), mood (*P* = .03), psychosis (*P* = .002) in 2020 compared to 2019 (Fig.1b).

Likewise, to determine if all diagnoses behave the same as the time changes (longitudinally), we have used a twoway ANOVA test for 2019 and 2020 separately. For 2019 and 2020, all the *P* -values are significant, meaning different diagnoses will have different intercepts and slopes for 2019 and 2020 (*P <* .001).

## Discussion

Herein we report changes in clinician diagnosed mental health disorders across age groups. Our findings are consistent with others who report an increase in mental health issues during the COVID-19 pandemic, especially for anxiety and depression ^4–6^. Specifically, we report an increase in anxiety, depression, bipolar disorder, mood disorder, and psychosis among individuals aged 26-35 during the lockdown phase of the COVID-19 pandemic in California.

The strength of our observations includes the “goldstandard” clinician diagnosis that is recorded in the electronic health record. Self-report surveys were not used. We investigated the longitudinal trends of mental health disorders during the same months before and during the pandemic to serve as a historical control, allowing us to to account for any seasonal effects often noted in mental health reports.

Additionally, because California implemented public health measures uniformly throughout the state, and there was not county-by-county differences in policies, EHR data facilities across the state can be used.

In short, those living in California would not have the benefit of learning from another state about the impact of a lockdown. Besides, due to the nature of the data set and the diversity of the state of California, this analysis captured the generalized statewide increase. California was the first to initiate a lockdown and, as such, would be the first to experience any sequelae from those safety measures.

Those ages 26-35 demonstrated the greatest increase in seeking care for a mental health disorder during the lockdown phase of the pandemic. There may be several explanations for this, including caring for and financial supporting a younger family, as well as stress and worry about loss of older family members such as parents and grandparents. Financial issues brought on by inability to work, change in employment status or cut backs from corporations or personal businesses may have contributed to the increase in anxiety and depression. The mental health disorders may have been further propelled by the increase in substance use observed in this age group, including an increase in self-harm behaviors and suicide. We also observed a reduction in many mental health disorders, especially among adults older than 55. The reason for this is unclear, but may be due to not seeking health care for fear of becoming infected with SARS-CoV-2, as it was widely publicized that advancing age and the presence of cardiovascular commodities increased risk of serious illness. Although our study utilized clinician diagnosis of mental health disorders there are limitations associated with the use of electronic health records. Primarily that all diagnoses were recorded and captured accurately, and that the data reported in the record only reflect that of individuals who have sought care. Furthermore, the UC-CORDs data set consists of record solely from academic health science centers which may not reflect trends in smaller communities or in areas where there are no academic health science centers.

Our observations demonstrate the impact of the lockdown and COVID-19 pandemic on mental health disorders in California. The use of public health measures, including the lock down, and the availability of vaccines has resulted in California having a COVID-19 death rate of 228 per 100,000, making it one of the lowest in the United States. However, we are just beginning to understand the impact of the totality of the pandemic and the measures instituted to stop the spread on mental and physical health. More studies are desperately needed to understand the factors that contribute to these issues so that interventions and policies can be instituted when another global pandemic emerges.

## Data Availability

All data produced in the present study are available upon reasonable request to the authors.

## Acknowledgements

The UC-CORDS data set, is made available by the University of California Office of the President and the University of California Biomedical, Research, Acceleration, Integration, and Development. Biomedical computing facilities are supported by the National Center for Research Resources and the National Center for Advancing Translational Sciences, National Institutes of Health, through Grant UL1 TR001414. The content is solely the responsibility of the authors and does not necessarily represent the official views of the NIH.

## Notes

### Competing Interest Statement

The authors have declared no competing interest.

### Funding Statement

This study did not receive any funding.

### Author Declarations

The use of the dataset was jointly reviewed by the Institutional Review Boards of all University of California Health campuses and was determined to be non-human subjects research.

